# Detection of parasite DNA in soil samples from rural Yucatán, México

**DOI:** 10.1101/2023.06.27.23291958

**Authors:** Liliana Villanueva-Lizama, Angela Cruz-Coral, Christian Teh-Poot, Vladimir Cruz-Chan, Rojelio Mejia

## Abstract

The soil is the primary environmental reservoir for many parasites transmitted to humans through the fecal-oral route, causing disease. Our environmental study used a high-throughput multi-parallel real-time quantitative PCR (qPCR) assay to detect parasites’ DNA in soil collected from the outdoor built environments of 34 houses. The total parasite prevalence was *Acanthamoeba spp*. (53%), *Blastocystis* spp. (12%), *Ascaris lumbricoides* (12%), *Toxocara canis* (9%), *Ancylostoma* spp. (3%), *Trichuris trichiura* (3%), *Entamoeba histolytica* (3%) and *Giardia intestinalis* (3%). No DNA from *Necator americanus, Strongyloides stercoralis, Toxocara catis*, and *Cryptosporidium* spp was detected. A total of 65% of houses were positive for at least one parasite, 15% had poly-parasites, and up to six different parasites were detected in a single sample. This is the first report of parasites-DNA detected in soil samples from a rural community in Yucatán and suggests higher rates of transmission of parasites with both a zoonotic and medical importance.

## SHORT COMMUNICATION

Some parasites present in the soil are an important health problem in tropical and subtropical regions. These parasitic infections are caused by soil-transmitted helminths (STH): *Ascaris lumbricoides, Ancylostoma duodenale, Necator americanus, Strongyloides stercoralis, Trichuris trichiura, Toxocara catis, Toxocara canis*, and protozoan parasites such as *Blastocystis* spp., *Entamoeba histolytica, Giardia intestinalis, Cryptosporidium* spp. Intestinal parasitic infections can cause diarrhea, malnutrition, anemia, impaired nutrition, development, and growth delays in children^1^. According to the World Health Organization (WHO), approximately 1.5 billion people (20% of the world’s population) are infected with STH worldwide, and its health impact is estimated at 5.2 million disability-adjusted life years (DALYs)^1^. Other soil-borne protists are opportunistic human pathogens. *Acanthamoeba* spp. are the most prevalent protozoa found in the environment^2^.

Parasites have relevance in environmental as well as human health. Soil contaminated with parasite eggs or cysts can favor the spread of the parasite to humans, and the rural communities have traditions that allow extensive contact with contaminated soil, such as eating/cooking outside, children playing in the yards, and using latrines. Therefore, assessing the outdoor built environment contamination can evaluate the risk of parasitic infections in rural communities. We aim to determine the presence of DNA from 12 different parasites, including helminths (*Ascaris lumbricoides, Ancylostoma* spp., *Necator americanus, Strongyloides stercoralis, Trichuris trichiura* and *Toxocara canis/cati);* the protozoa include *(Cryptosporidium* spp., *Entamoeba histolytica, Acanthamoeba* spp., *and Giardia intestinalis);* and heterokonts *(Blastocystis* spp.) in soil collected from a rural community in Yucatan, using a previously standardized multi-parallel real-time quantitative PCR. This study aims to incorporate the relevance of several zoonotic and human parasites and the risk of infection to populations living in endemic regions.

The study was performed in Sudzal, a rural Yucatán, México community with about 1700 inhabitants. This community has traditional customs of latrine use and eating/cooking outside habits. Soil samples were collected from the outdoor build environment of 34 households randomly selected after obtaining consent from the owners. Samples were collected per duplicate in 50 mL Falcon tubes (containing approximately 50 mg of wet soil). A total of 68 samples were stored at -20ºC until use. Samples were weighed and resuspended in 35.6% w/w sodium nitrate in water and centrifuged at 1500 x g for 5 min. Supernatants were filtered through a 3.0 µm SSWP membrane (MF-Millipore, Merck KGaA, Darmstadt, Alemania), and the filtered materials were processed using FastDNA SPIN kit for soils (MP Biomedicals, Santa Ana, California, USA) as previously described^3^. DNA eluent was dried on 0.2 µm filter paper (Millipore, Merck KGaA, Darmstadt, Alemania) and shipped to Baylor College of Medicine Laboratories at ambient temperatures. DNA was extracted from these filter papers using 100 µl of DEC buffer (MP Biomedicals). DNA from soil samples was analyzed by a multi-parallel quantitative polymerase chain reaction (qPCR) as described by Mejia et al. ^4^. In addition, *Acanthamoeba* DNA was detected using a set of primers and probes previously reported^5^.

Forward Primer: CCCAGATCGTTTACCGTGAA

Reverse Primer: TAAATATTAATGCCCCCAACTATCC

Probe 5’FAM MGB: CTGCCACCGAATACATTAGCATGG

Samples were run on a ViiA 7 Fast Real-time PCR System (Applied Biosystems, Waltham, Massachusetts, USA), and plasmids containing the target sequences were run duplicated to create a standard curve. An exogenous internal control was used to assess the efficiency of DNA extraction, as described previously^4,6^.

Of the 34 houses tested, 22 (65%) were positive for the DNA presence of at least one parasite, and 12 (35%) were negative. Up to six different parasites were detected in a single sample, and 15% of the samples had more than one parasite detected. Over fifty percent (56%) of the samples analyzed were positive for at least one protozoon, and 18% were positive for at least one helminth parasite species. Three out of 22 (14%) samples were positive for protozoa and helminth parasites. Meanwhile, 19/22 (86%) were samples contaminated only with protozoa and 3/22 (14%) with helminth species. *Acanthamoeba* spp. had the highest DNA concentration compared to the other parasites (P < 0.0001) (Fig 1).

**Fig. 1.**
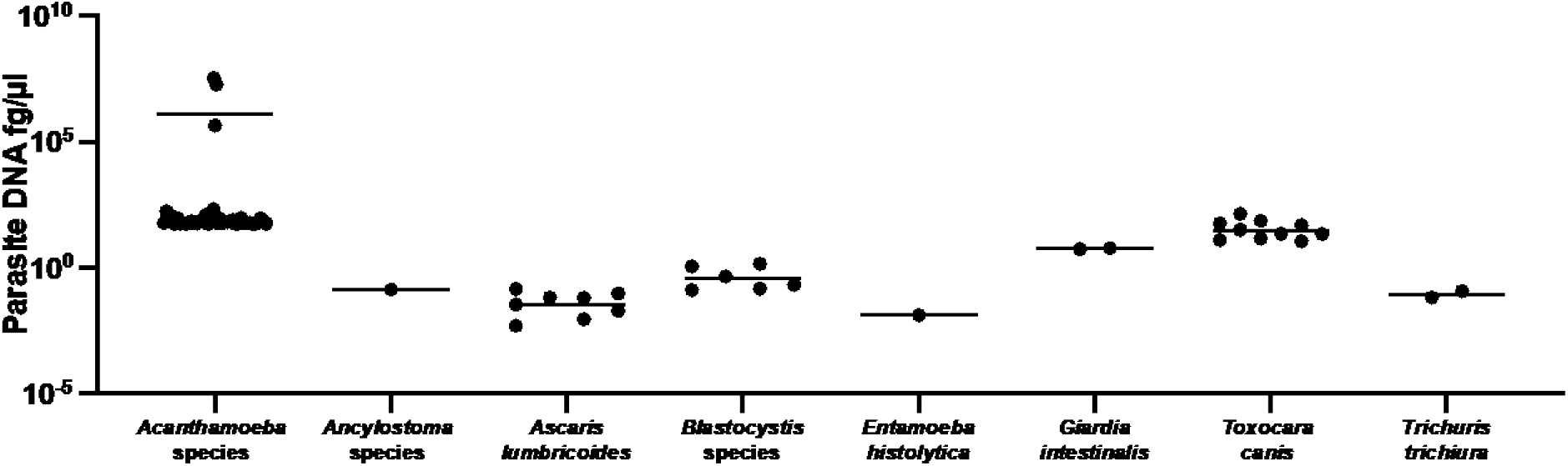
Total prevalence of parasites in all 68 samples. *Acanthamoeba* spp. had the highest DNA concentration compared to all other parasites (Kruskal Wallis P < 0.0001).

Multi-parallel qPCR revealed that *Acanthamoeba* spp was the most frequent protozoa (18/34, 53%), and *Ascaris lumbricoides* was the most frequent helminth (4/34, 12%) detected. Positivity rates and DNA concentration range detected per parasite specie are presented in Table 2: *A. lumbricoides* (12%), *T. canis (9%), A. duodenale, T. trichiura*, and *E. histolytica* (3% each). *Blastocystis* was detected in 12% of the samples. No positive samples were detected for *N. americanus, S. stercoralis, T. cati*, or *Cryptosporidium*.

**Table 1.**
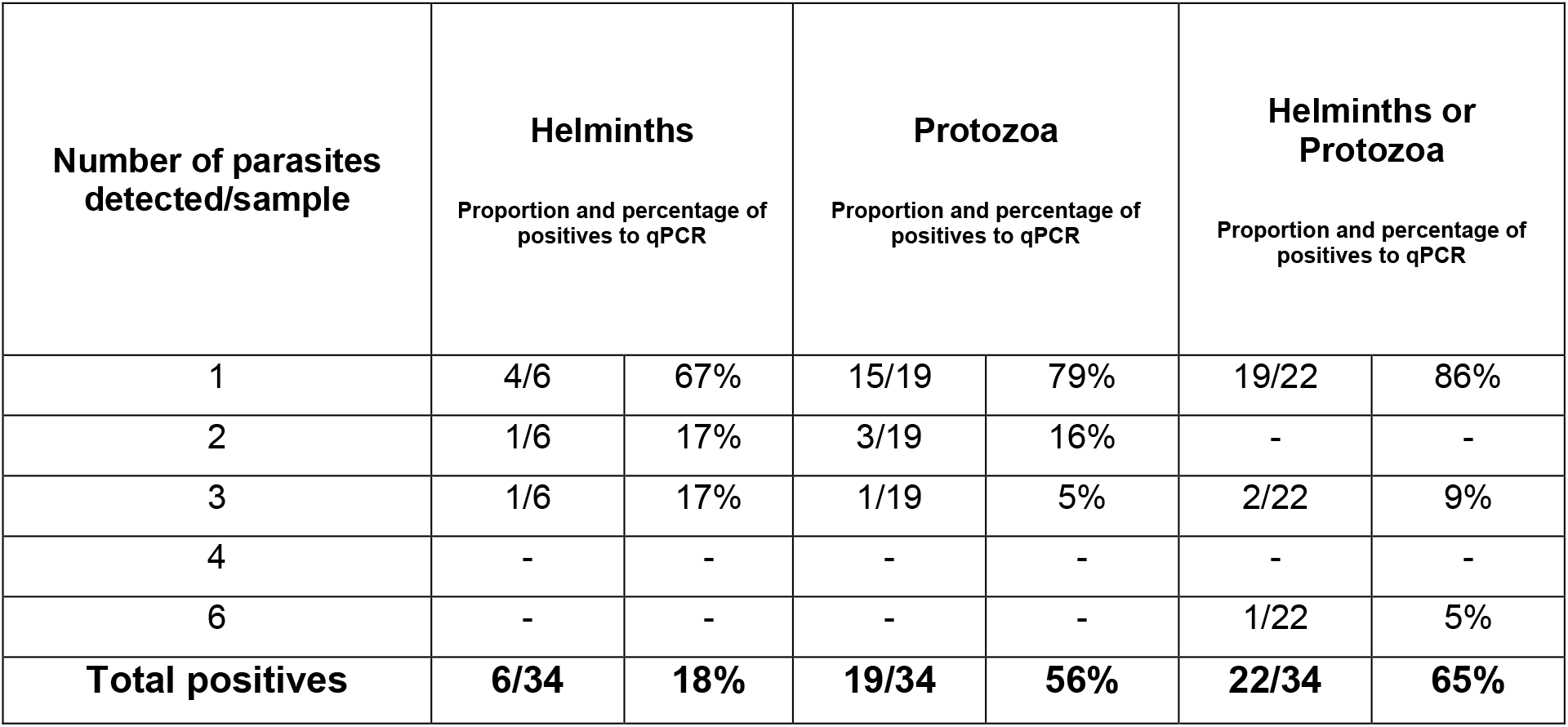
Proportion and percentage of samples positive to Helminths or Protozoa among the study population.

**Table 2.** Percentage of positives and DNA concentration measured by qPCR in the soil samples collected.

|  | % PCR positives | DNA concentration in soil<br>(fg/μl, median (range)) |
| --- | --- | --- |
| *Acanthamoeba* spp. | 53%, (18/34) | 63.3373 (54.95 - 3447) |
| *Ascaris lumbricoides* | 12%, (4/34) | 0.035 (0.005-0.144) |
| *Ancylostoma* spp. | 3%, (1/34) | 0.135 |
| *Blastocystis* spp. | 12%, (4/34) | 0.331 (0.13-1.272) |
| *Cryptosporidium* spp. | 0%, (0/34) | 0 |
| *Entamoeba histolytica* | 3%, (1/34) | 0.013 |
| *Giardia intestinalis* | 3%, (1/34) | 6.087 |
| *Necator americanus* | 0%, (0/34) | 0 |
| *Strongyloides stercoralis* | 0%, (0/34) | 0 |
| *Trichuris trichiura* | 3%, (1/34) | 0.091 |
| *Toxocara canis* | 9%, (3/34) | 27.16689038 (11.43-137.245) |
| *Toxocara cati* | 0%, (0/34) | 0 |

To our knowledge, this is the first study reporting the molecular detection of *Acanthamoeba* spp. in soil samples from Yucatán. *Acanthamoeba* is an amphizoid organism that is a free-living amoeba in nature and a parasitic pathogen^2^. *Acanthamoeba* is the causative agents of granulomatous amebic encephalitis (GAE), a potentially fatal disease of the central nervous system, amebic keratitis (AK), a painful disease of the eyes, cutaneous lesions, and sinusitis in immunocompromised patients^7^. More studies should be performed to identify the *Acanthamoeba* species present in the soil collected in rural Yucatan and identify the risk to human health and its participation in the ecology of the environment.

*T. catis* and *T. canis* are helminthic worms that infect cats and dogs. Both animals can defecate thousands of eggs into the environment. DNA from *T. canis* was detected in the soil sampling collected in our study. Previous studies performed in fecal samples from dogs belonging to rural communities from Yucatán reported a frequency of 6% (8/130) of *T. canis*^8^. Sampling collection for this study includes soil collected from areas where children play, suggesting environmental contamination and risk for human toxocariasis. Our results indicate that feral and domestic dogs in Sudzal represent a significant source of environmental contamination. Other studies have reported similar frequencies of *T. canis* eggs and DNA in public spaces from poorer neighborhoods, showing an association of toxocariasis with poverty conditions^9^.

Panti-May et al.^10^ studied the prevalence of intestinal parasites in children from rural Yucatán and reported an overall majority of 65%; six protozoa and four helminths were identified using two coproparasitological tests. *Blastocystis* spp (44.6%), *Giardia intestinalis* (26.5%), and *Entamoeba coli* (26.5%) were the most prevalent parasites detected. *A. lumbricoides, T. trichiura*, and *B. hominis* were also detected in children, showing the need to perform diagnostics for intestinal parasites in children and dogs living in rural Yucatán.

A few studies have previously reported soil contaminated with parasite eggs in rural and urban settings, with prevalence ranging from 11% to 82%, suggesting differences due to geography^11^. However, these studies have used microscopy techniques for egg counting and detection. Detection of parasite eggs in environmental samples such as soil or feces requires traditional techniques such as flotation/microscopy, although it is well known that microscopy’s sensitivity is very low^12^. DNA-detection-based tools represent an alternative to parasitological methods for accurate diagnostics, monitoring worm burden before and after treatment, detecting genetic markers related to anthelmintic resistance, or monitoring drug-efficacy trials^13^. Quantitative polymerase chain reaction (qPCR)-based assays represent a promising alternative approach for the diagnosis of infection or epidemiological research purposes. Although more expensive than standard microscopy, molecular diagnostic is already used in many low and middle-income countries due to its sensibility and specificity, more accurate quantification, increased throughput, requiring fewer trained personnel, and offering insight into the relationship among multiple pathogens^14^. The method used here to identify the presence/contamination of soil with parasites is more sensitive than microscopy methods.

The limitations of this study include a small sample size for enrolled houses, thus not giving a broader view of the parasite prevalence in the region. Also, only two samples were collected per built environment, limiting the understanding of these parasites’ locations. Only one *Ancyslostoma* helminth was detected, but the primer and probes are specific to the genus, and unclear if this is an animal or human pathogen.

Finally, our findings suggest soil contamination is present in the outdoor built environment of households in rural Yucatán, Mexico. Detection of parasite DNA in the soil of above 50% of the houses evaluated poses a health risk for the inhabitants. Moreover, 15% of the houses assessed had soil contaminated with more than one parasite species, and up to 6 different parasite species were detected in a single home. This is the first report of parasites-DNA detected in soil samples from a rural community in Yucatán and suggests higher transmission rates of parasites with medical importance. Furthermore, an easy-to-use qPCR identified several parasites critical to decision-making for treatment and public health interventions.

## Data Availability

All data produced in the present study are available upon reasonable request to the authors

## Acknowledgments

The authors would like to thank the people of Yucatán, Mexico for welcoming us into their homes.

## REFERENCES

1. Soil-transmitted helminth infections. World Health Organization. Accesed May 9, 2023. https://www.who.int/news-room/fact-sheets/detail/soil-transmitted-helminth-infections

2. Geisen S, Mitchell EAD, Adl S, et al. Soil protists: a fertile frontier in soil biology research. FEMS Microbiol Rev. 2018;42(3):293–323. doi:10.1093/femsre/fuy006

3. Mejia R, Seco-Hidalgo V, Garcia-Ramon D, et al. Detection of enteric parasite DNA in household and bed dust samples: potential for infection transmission. Parasites – vectors. 2020;13(1),141. doi.org/10.1186/s13071-020-04012-6

4. Mejia R, Vicuña Y, Broncano N, et al. A novel, multi-parallel, real-time polymerase chain reaction approach for eight gastrointestinal parasites provides improved diagnostic capabilities to resource-limited at-risk populations. Am J Trop Med Hyg. 2013;88(6):1041–1047. doi:10.4269/ajtmh.12-0726

5. Aykur M., and Dagci H. Evaluation of molecular characterization and phylogeny for quantification of Acanthamoeba and Naegleria fowleri in various water sources, Turkey. PloS one. 2021;16(8),e0256659. doi.org/10.1371/journal.pone.0256659

6. Naceanceno KS, Matamoros G, Gabrie JA, et al. Use of Multi-Parallel Real-Time Quantitative PCR to Determine Blastocystis Prevalence and association with other gastrointestinal parasite infection in a rural Honduran location. Am J Trop Med Hyg. 2020;102(6):1373–1375. doi:10.4269/ajtmh.19-0876

7. Marciano-Cabral F, Cabral G. Acanthamoeba spp. as agents of disease in humans. Clin Microbiol Rev. 2003;16(2):273–307. doi:10.1128/CMR.16.2.273-307.2003

8. Rodríguez-Vivas RI, Gutierrez-Ruiz E, Bolio-González ME, et al. An epidemiological study of intestinal parasites of dogs from Yucatan, Mexico, and their risk to public health. Vector Borne Zoonotic Dis. 2011;11(8):1141–1144. doi:10.1089/vbz.2010.0232

9. Tyungu DL, McCormick D, Lau CL, et al. Toxocara species environmental contamination of public spaces in New York City. PLoS Negl Trop Dis. 2020;14(5):e0008249. doi:10.1371/journal.pntd.0008249

10. Panti-May JA, Zonta ML, Cociancic P, et al. Occurrence of intestinal parasites in Mayan children from Yucatán, Mexico. Acta Trop. 2019;195:58–61. doi:10.1016/j.actatropica.2019.04.023

11. Steinbaum L, Njenga SM, Kihara J, et al. Soil-transmitted helminth eggs are present in soil at multiple locations within households in rural Kenya. PLoS One. 2016;11(6):e0157780. doi:10.1371/journal.pone.0157780

12. Mbong Ngwese M, Prince Manouana G, Nguema Moure PA, et al. Diagnostic techniques of soil-transmitted helminths: Impact on control measures. Trop Med Infect Dis. 2020;5(2). doi:10.3390/tropicalmed5020093

13. Gandasegui J, Martínez-Valladares M, Grau-Pujol B, et al. Role of DNA-detection-based tools for monitoring the soil-transmitted helminth treatment response in drug-efficacy trials. PLoS Negl Trop Dis. 2020;14(2):e0007931. doi:10.1371/journal.pntd.0007931

14. Easton A V, Oliveira RG, O’Connell EM, et al. Multi-parallel qPCR provides increased sensitivity and diagnostic breadth for gastrointestinal parasites of humans: field-based inferences on the impact of mass deworming. Parasit Vectors. 2016;9:38. doi:10.1186/s13071-016-1314-y

